# Enrichment for Cases of African-American Patients with Pathogenic *TTR* V142I Variant in the TOPCAT Trial

**DOI:** 10.1101/2020.10.14.20201046

**Authors:** Nikolaos Papoutsidakis, Neeru Gandotra, Edward Miller, Michael F. Murray, Curt Scharfe, Daniel Jacoby

**Affiliations:** Department of Internal Medicine, Cardiovascular Medicine, Yale School of Medicine, New Haven, CT; Department of Genetics, Yale School of Medicine, New Haven, CT

## Abstract

Transthyretin cardiac amyloidosis (ATTR-CA) is a treatable cause of heart failure with a hereditary form that disproportionally affects patients of West African ancestry. The clinical management of ATTR-CA has dramatically changed in the past five years, with rapidly evolving diagnostic approaches and life-prolonging therapies. The *TTR* variant c.424G>A, p.V142I (aka V122I) is pathogenic and occurs in 3-4% of individuals of West African ancestry. Despite its high frequency, V142I ATTR-CA is often unrecognized due to variable clinical penetrance, limited knowledge, and lack of inexpensive non-invasive diagnostic tests. Currently unknown is which *TTR* V142I carriers will progress to heart failure and at what age. Here we studied the prevalence of *TTR* V142I among a random cohort of African-American patients enrolled in the Treatment of Preserved Cardiac Function Heart Failure With an Aldosterone Antagonist Trial (TOPCAT). Three of the 26 HFpEF patients (11.5%) studied carried the pathogenic *TTR* V142I variant. While we cannot conclude at this point that *TTR* V142I was the underlying cause of the clinical phenotype in these patients, our results suggest that rapid *TTR* V142I genotyping, in combination with heart imaging, could have immediate clinical utility for identifying under-/mis-diagnosed HFpEF patients.

## Introduction

Transthyretin cardiac amyloidosis (ATTR-CA) is an increasingly recognized cause of left ventricular hypertrophy and heart failure. Pathogenic DNA variants in the *TTR* gene confer an autosomal dominant risk for ATTR, with clinical manifestations that precede diagnosis often by years[1]. One of the most frequent pathogenic *TTR* variants (c.424G>A, p.V142I, also notated as V122I) occurs almost exclusively in individuals of West African descent[2]. An estimated 1.5 million African Americans (3.4%) in the United States are heterozygous for *TTR* V142I[2]. Heart failure with preserved ejection fraction (HFpEF) is a heterogeneous clinical phenotype that accounts for approximately half of newly diagnosed cases of heart failure in the US per year and for which there are currently no evidence based medical therapies. Recent studies suggested that ATTR cardiomyopathy prevalence amongst HFpEF patients is much higher than previously thought, due to under-recognition[2, 3]. This has important implications for HFpEF clinical trials as treatment effects are likely to differ between idiopathic and ATTR-related HFpEF. Here we describe our investigation of the prevalence of *TTR* V142I among a random cohort of African-American patients enrolled in the Treatment of Preserved Cardiac Function Heart Failure With an Aldosterone Antagonist Trial (TOPCAT)[4].

## Material and Methods

We requested all available trial data and de-identified DNA samples from patients enrolled in TOPCAT who self-identified as black from the BIOLINCC repository. *TTR* gene variants were identified using clinical exome sequencing and confirmed using bidirectional Sanger sequencing. The genetic findings were correlated with clinical outcome data for each subject. All but one of the samples originated from US sites. This study was overseen by the institutional review board at Yale University.

## Results

The BIOLINCC repository was able to provide biospecimens with extractable DNA for only 26 TOPCAT black subjects, possibly due to elapsed time from the TOPCAT trial. Three of 26 HFpEF patients studied carried the pathogenic *TTR* V142I variant (Table 1). The enrichment of *TTR* V142I in this cohort of HFpEF patients (11.5%) was increased compared to the general African-American population (3.4%). Analysis of trial data showed that all three subjects positive for *TTR* V142I were assigned in the placebo group, and two out of three patients (66%) reached the primary end-point (composite of death from cardiovascular causes, aborted cardiac arrest, or hospitalization for the management of heart failure). In comparison, of the 302 black TOPCAT patients for whom trial data were available, 34.4% (n=104) reached the primary end-point.

**Table 1:** Demographic and clinical data from 26 TOPCAT patients and genetic findings.

|  |  |  |  |  |  |  |  |  |  |  |  |  |  |
| --- | --- | --- | --- | --- | --- | --- | --- | --- | --- | --- | --- | --- | --- |
| Random ID | 710811 | 875425 | 343485 | 220741 | 948460 | 233830 | 487405 | 990265 | 886315 | 415485 | 313427 | 476332 | 988134 |
| Pathogenic TTR Variant | c.424G>A (p.V142I) | c.424G>A (p.V142I) | c.424G>A (p.V142I) | - | - | - | - | - | - | - | - | - | - |
| Gender | F | M | F | F | F | F | F | M | M | F | M | F | F |
| Age Entering Study | 70's | 60's | 50's | 80's | 60's | 80's | 60's | 60's | 70's | 60's | 50's | 60's | 50's |
| TOPCAT group* | PCB | PCB | PCB | SP | PCB | PCB | PCB | PCB | SP | SP | SP | PCB | PCB |
| Primary Endpoint reached | N | Y | Y | N | Y | Y | Y | Y | Y | N | N | N | N |
| Any Cause death | N | N | N | N | Y | N | N | Y | N | N | N | N | N |
| SBP (mm Hg) | 160 | 118 | 133 | 126 | 120 | 141 | 136 | 122 | 131 | 150 | 126 | 97 | 118 |
| DBP (mm Hg) | 52 | 75 | 71 | 70 | 69 | 69 | 72 | 82 | 61 | 58 | 88 | 65 | 60 |
| Use of BB | Y | Y | Y | Y | Y | Y | Y | Y | Y | Y | Y | Y | Y |
| Use of CCB | Y | Y | N | Y | N | N | Y | Y | N | Y | Y | N | Y |
| Use of Diuretic | Y | Y | Y | Y | Y | Y | Y | Y | Y | Y | Y | Y | Y |
| Use of ACE-I or ARB | Y | Y | Y | N | N | Y | Y | Y | Y | N | Y | Y | Y |
| LVH on ECG | Y | N | N |  | N | Y | N | N | N | N | N |  | N |
| Interventricular septum thickness | - | - | 1.1 | 0.9 | - | 1.2 | 1.2 | - | 1.3 | 1.2 | 1.1 | 1.1 | - |
| Posterior wall thickness | - | - | 1.1 | 0.9 | - | 1.1 | 1.1 | - | 1.1 | 1.2 | 1.1 | 1.0 | - |
| LV Mass | - | - | 197 | 166 | - | 179 | 245 | - | 213 | 272 | 154 | 217 | - |
| Ejection Fraction (prior to enrolling) | 60 | 50 | 52 | 60 | 70 | 60 | 76 | 50 | 45 | 60 | 45 | 50 | 48 |
| Longitudinal Strain | - | - | -13.29 | -18.61 | - | - | -15.62 | - | - | -18.28 | -10.39 | -18.63 | - |
| Hypertension | Y | Y | Y | Y | Y | Y | Y | Y | Y | Y | Y | Y | Y |
| Atrial fibrillation | N | Y | Y | N | N | Y | N | N | Y | N | Y | N | N |

\* PCB=Placebo, SP=Spironolactone

**Table 1** (continued)
|  |  |  |  |  |  |  |  |  |  |  |  |  |  |
| --- | --- | --- | --- | --- | --- | --- | --- | --- | --- | --- | --- | --- | --- |
| Random ID | 954421 | 689847 | 437794 | 386230 | 302130 | 637663 | 442800 | 555747 | 371036 | 715887 | 955499 | 908592 | 339794 |
| Pathogenic TTR Variant | - | - | - | - | - | - | - | - | - | - | - | - | - |
| Gender | F | F | F | F | M | F | F | F | F | F | M | F | F |
| Age Entering Study | 60's | 60's | 50's | 60's | 60's | 70's | 60's | 70's | 50's | 60's | 50's | 60's | 60's |
| TOPCAT group* | SP | SP | PCB | SP | SP | SP | PCB | SP | PCB | PCB | PCB | SP | SP |
| Primary Endpoint reached | N | Y | N | Y | Y | Y | Y | Y | Y | Y | Y | Y | N |
| Any Cause death | N | N | N | N | Y | N | N | N | N | Y | N | N | N |
| SBP (mm Hg) | 143 | 138 | 133 | 135 | 146 | 136 | 147 | 124 | 110 | 119 | 121 | 154 | 128 |
| DBP (mm Hg) | 73 | 70 | 92 | 90 | 90 | 58 | 85 | 70 | 64 | 63 | 85 | 96 | 58 |
| Use of BB | Y | Y | Y | N | Y | Y | Y | Y | Y | Y | Y | Y | Y |
| Use of CCB | Y | Y | Y | N | N | Y | Y | N | Y | Y | N | Y | Y |
| Use of Diuretic | Y | Y | Y | Y | Y | Y | Y | Y | Y | Y | Y | Y | Y |
| Use of ACE-I or ARB | Y | N | Y | Y | N | Y | Y | N | Y | Y | Y | Y | Y |
| LVH on ECG | N | N | Y | Y | N | Y | N | N | N | Y | N | - | N |
| Interventricular septum thickness | 1.4 | 1.2 | 1.6 | 1.3 | 1.4 | - | - | 1.1 | - | - | - | 1.5 | - |
| Posterior wall thickness | 1.4 | 1.1 | 1.6 | 1.2 | 1.5 | - | - | 1.1 | - | - | - | 1.3 | - |
| LV Mass | 317 | 141 | 276 | 227 | 336 | - | - | 198 | - | - | - | 321 | - |
| Ejection Fraction (prior to enrolling) | 60 | 72 | 50 | 45 | 45 | 67 | 58 | 65 | - | - | - | - | - |
| Longitudinal Strain | -17.83 | -19.43 | -12.96 | -13.50 | -13.62 | - | - | -13.21 | - | - | - | - | - |
| Hypertension | Y | Y | Y | Y | Y | Y | Y | Y | Y | Y | Y | Y | Y |
| Atrial fibrillation | N | N | N | N | N | N | N | Y | N | N | N | Y | N |

\* PCB=Placebo, SP=Spironolactone

## Discussion

We found a significant proportion of African-American HFpEF subjects (11.5%) carried the *TTR* V142I variant in this random sample from TOPCAT, in spite of efforts to specifically exclude infiltrative cardiomyopathy at the point of enrollment[4]. While we cannot say for certain, it is possible that these three patients suffered from HFpEF driven by ATTR-CA at the time of enrollment. ATTR-CA runs an insipid course, with TTR amyloid tissue deposition often preceding heart failure diagnosis by years[1, 2]. Our findings are consistent with prior findings suggesting that ATTR-CA can be missed even after current approaches to strict clinical trial screening. Our sample cohort was significantly enriched in females, for unknown reasons. This could potentially alter patients’ clinical characteristics, as inherited ATTR-CA often expresses at a later age in females. Whether enrolling unidentified ATTR-CA patients in clinical HFpEF trials adversely affects our ability to identify effective treatments is a relevant question.

There has been a series of negative HFpEF trials over the years leading to near complete absence of effective medical treatment for HFpEF. Could it be possible that occult ATTR-CA has been one of the drivers of the extended HFpEF therapy drought? That depends on whether we believe that ATTR-CA patients would have a different response to medical therapy than “run-of-the-mill” HFpEF. There is general agreement that angiotensin pathway modification and beta-blockade are at best ineffective and at worst counterproductive in the treatment of patients with ATTR-CA. Whether aldosterone receptor antagonists (studied in TOPCAT) react differently is at present impossible to say. In any event, ATTR screening was not part of previous studies and merits consideration for HFpEF and, potentially, even HFrEF[5] clinical trials going forward. For populations shown to be enriched with ATTR (African-Americans middle-aged or older, or elderly men regardless of race with HFpEF) we would contend that it is worthwhile to explore point-of-care diagnostic strategies, employing 99mTc-pyrophosphate (PYP) cardiac scanning and/or rapid *TTR* V142I genetic testing when enrolling for clinical trials for HFpEF.

## Data Availability

All data referred to in the manuscript are provided in Table 1.

